# Are There Sex Differences in Thrombectomy Utilisation in Treatment of Acute Ischaemic Stroke? A Systematic Review and meta-analysis

**DOI:** 10.1101/2023.03.11.23287149

**Authors:** Serena Baker, Clayton Micallef, Gillian Mead

## Abstract

**Objectives:** Mechanical thrombectomy (MT) is a highly effective treatment for acute ischaemic stroke (AIS). Our aim was to determine whether there are differences in access to thrombectomy between men and women; this is an important question because a previous meta-analysis had shown that women were less likely than men to receive intravenous thrombolysis for AIS.

**Materials:** This was a systematic review and meta-analysis.

**Methods:** Medical databases (Embase, Medline and APA Web of Science) were searched for eligible studies from 01/01/2010-30/09/2021. Two independent authors screened titles and abstracts and scrutinised full texts. Eligible studies were hospital-based, registry-based, or administrative data studies reporting sex-specific data on patients treated with thrombectomy for AIS, in representative populations of patients with AIS. Studies including only posterior circulation strokes were excluded. Summary unadjusted odds ratios were calculated to compare MT utilisation in men and women.

**Results:** 1,379 citations were retrieved, 76 underwent full review and 16 were included in the meta-analysis, which comprised 5,281,009 stroke cases (47.2% women, 52.8% men). The summary unadjusted OR for sex differences in thrombectomy use was 0.931 (95% CI 0.834-1.040, p=0.206), indicating women had lower odds of receiving MT though confidence intervals overlapped one. There was statistically significant heterogeneity between studies (Q=1043.13 p<0.0001, I^2^=98.56%).

**Conclusion:** We found no clear evidence that women were less likely to receive thrombectomy then men. Future studies should continue to report sex-specific data to ensure that there is equity of access to thrombectomy irrespective of sex.

## Introduction

Acute ischaemic stroke (AIS) represents a significant burden of mortality and morbidity globally, representing approximately 11% of all deaths in 2019, second only to ischaemic heart disease^1^. The lifetime risk of AIS does not differ according to sex^2^; but whether differences exist in the presentation, treatment and outcome of AIS between men and women is debated in literature.

A person’s sex refers to their biological characteristics, usually categorised as “male” or “female” after birth. Gender is a person’s expression of self, representing socially constructed roles and behaviours^3^. Neither are binary, and they can be non-concordant. It is difficult to differentiate sex and gender in stroke literature, therefore this review discusses sex as “male” or “female” while noting potential inaccuracies, and oversight of non-conforming patients.

Intra-venous thrombolysis (IVT, tissue plasminogen activator) is an AIS therapy that can be given within 4.5 hours of symptom onset^4^. Two large meta-analyses using data from 2000-2018 found that women were less likely to receive IVT than men. This difference was larger in 2000-2008 than between 2008-2018, yet women were still 13% less likely to be given IVT. The difference may partially be explained by women being older and presenting later after symptom onset, both factors affecting their eligibility for IVT^5^, though age should not determine access, as older patients benefit equally from IVT^6^.

Mechanical thrombectomy (MT) is a treatment for AIS is becoming more widely used after strong evidence of its benefit was published in 2015^7-11^. MT is used to treat proximal anterior circulation occlusions (PACO) and can be given up to 24 hours after symptom onset, a benefit over IVT. Importantly, rates of intra-cerebral haemorrhage, a side effect of IVT, did not increase with MT treatment. MT is equally effective in men and women, and in patients over 80 years old^12^. Therefore, women, even though they may present later after symptom onset and have strokes at an older age, should benefit from MT treatment.

As MT is equally effective in both sexes^12^, there should, be no difference in rates of MT utilisation. But given the sex differences in IVT use, it is important to determine whether sex differences exist for MT too. Stroke care must be equitable and protected characteristics of patients, including sex, gender, age, race and ethnicity^13^, should not affect care.

The aim of this systematic review was to analyse MT utilisation for treatment of AIS to examine if sex differences exist and explore rates of MT use according to potential confounding factors.

## Methods

This protocol was published in PROSPERO (registration number CRD42021277302). This methodology is based on a published meta-analysis of sex differences in IVT use^5^.

### Eligibility Criteria and Search

Embase, Medline and American Psychological Association (APA) Web of Science databases were searched between 01/01/2010-30/09/2021 using the following combination of terms: (1) “stroke” or “cerebrovascular accident” or “cerebral infarct”; (2) “sex” or “sex difference” or “sex distribution” or “sex factor”; and (3) “thrombectomy” or “endovascular therapy” or “endovascular procedure” or “endovascular treatment”. Guidance on conducting systematic reviews was found in PRISMA guidelines^14^.

Relevant references were observational studies reporting sex-specific treatment rates for AIS in typical hospital settings, including specialist stroke centres, regional hospitals, hospital systems or hospital-based stroke registries. Conference abstracts, other grey literature and non-English texts were excluded. An ad hoc decision was made to exclude studies using only posterior circulation stroke, as insufficient evidence for MT benefit in this clinical scenario exists^15^. Studies analysing all AIS admissions were included, therefore a small number of posterior circulation strokes may be included in this analysis.

### Study Selection and Data Abstraction

Citations were imported into COVIDENCE. Two authors (SB, CM). screened titles and abstracts independently using COVIDENCE Full texts of potentially relevant references were obtained, and inclusion/exclusion criteria applied independently by two reviewers. Disagreement on eligibility was discussed with a third author (GM). Studies that met eligibility criteria had data abstraction completed independently using COVIDENCE. Pilot data extraction was carried out on two papers to assess the protocol.

We categorised study design as: hospital-based, registry-based, or administrative data. To differentiate between registries and administrative data, unless a study specified the use of a registry, it was categorised as administrative data. Extracted data describing study population: mean age and range, sex proportion, proportion of minority subjects (based on reported ethnicity). Other extracted data: study design, time period of enrolment, size of numerator and denominator populations, sex-specific treatment rates (%), odds ratios (ORs) describing sex difference (unadjusted and adjusted), 95% confidence intervals (CIs) and confounders that were adjusted for.

Ideally, all of the included papers would have used data from separate databases, or from the same database at separate time points. Two studies^16,17^ (n=572,395) used the same database with an overlap of three months, but a post-hoc decision was made to include both studies in the final analysis as the overlap was short compared to the overall length of both studies (12 months^16^ and 15 months^17^).

### Quality Scoring

Two authors (SB,CM) independently carried out quality assessment using a system adapted from the Newcastle-Ottawa Scale^18^ which has four criteria:

1. Representativeness of overall study population

2. Number and impact of exclusions applied to the initial patient cohort

3. Adjustment for potential confounding variables that might affect sex differences in MT use

4. Method by which outcome was ascertained

Each reference is given a score of 0-2 for each criterion where 2 is the best and 0 is the worst. An aggregated score was given on a 0-8 scale. This method of quality scoring was based on a published meta-analysis of sex differences in IVT use^5^. Examples of quality scoring using this technique given in supplementary material.

An additional quality score was given, based on a published meta-analysis of sex differences in IVT use^5^. Studies were graded A-D based on the primary focus. These categories were defined in advance of quality scoring. Group A papers had a clear objective to study sex differences in MT use. Group B papers examined sex differences in MT effectiveness but have no clear objective to study differences in use. Group C studies focus on MT treatment more broadly with no specific objective to look at sex difference. Group D studies focused on sex differences in acute stroke care with no objective to study MT use. This classification was developed as group A studies may have more validity for examining sex differences in MT use, when compared to group D studies.

### Statistical Analysis

Primary outcome measure of interest was the unadjusted OR comparing MT treatment rate in women with men in all AIS admissions. If not reported, this was calculated using MedCalc software^19^. Random effects-based results were reported due to variability in study design. Heterogeneity between studies was assessed using the Cochrane Q statistic and the I^2^ statistic. Where reported, we also extracted ORs adjusted for clinical characteristics. We had intended to perform an analysis of adjusted OR providing there were sufficient studies reporting these. Prespecified subgroup analyses (reported in PROSPERO protocol) include geographic region (North America, Europe, and Asia), study design (hospital-based studies and registries vs administrative data), quality score (0-8), and primary focus (A-D).

### Ethical Considerations

Ethical review board approval and informed consent was not required as this study made use of de-identified, published data.

## Results

The initial search yielded 1,379 citations; 76 potentially relevant full texts were obtained; (Figure 1); of these, 16 met eligibility criteria and provided sufficient sex-specific data to calculate an unadjusted OR comparing sex-specific MT rates. There were a total of 5,281,009 stroke patients (2,490,698 women, 47.2%). Study characteristics and MT treatment rates by sex are summarised in Table1. Throughout all of the studies, 69,838 women (2.80%) and 71,482 men (2.56%) were treated with MT.

**Figure 1:**
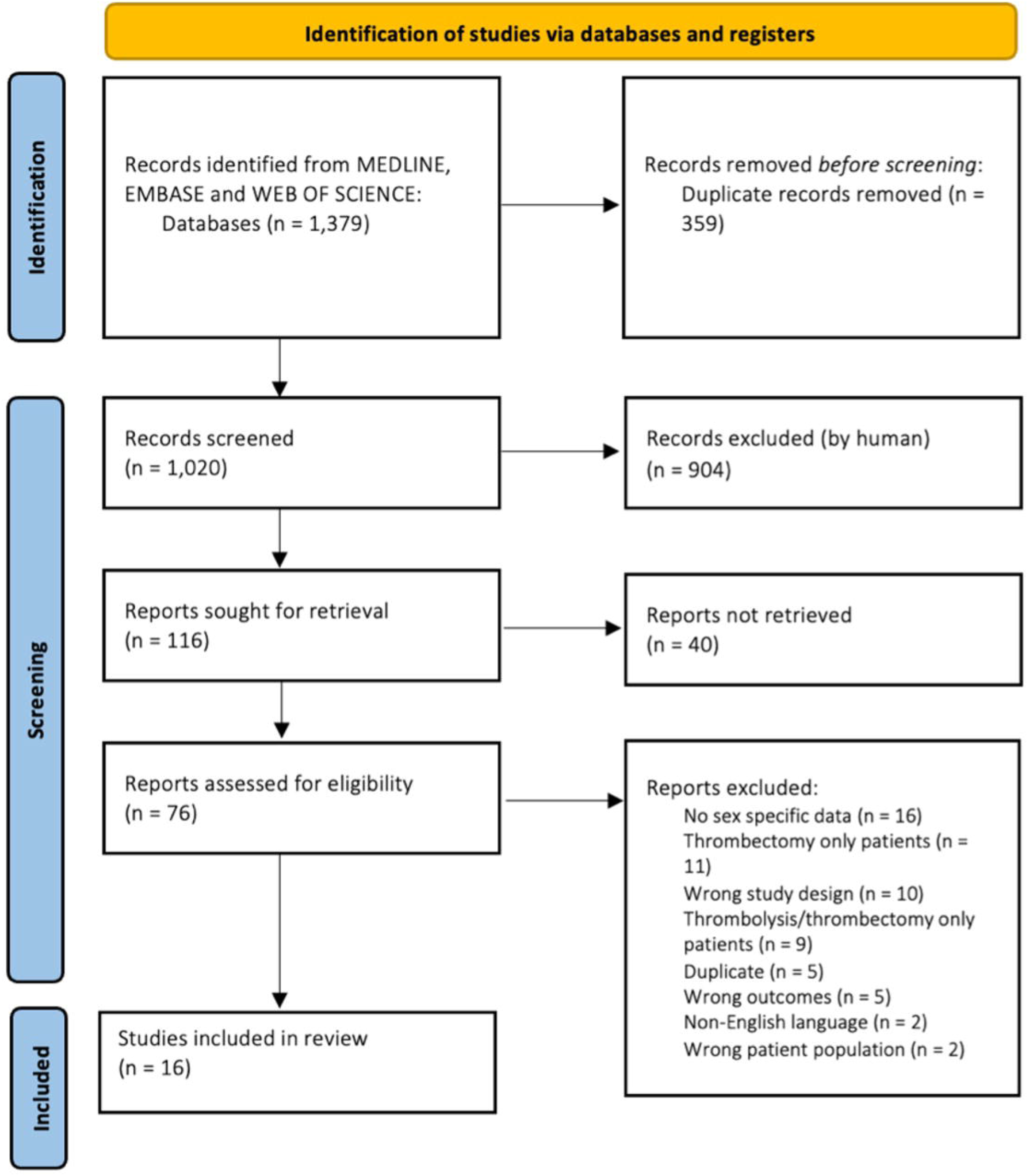
PRISMA flow diagram^20^ providing reasons for exclusion of full-text studies

There were two hospital-based studies, six registry-based and ten studies using administrative data. Study size varied from 169 patients to 1,443,894 and MT treatment rates ranged from 0.26%-50.77% Quality scores of studies varied from 4 to 7, with 14 studies scoring either 5 or 6. Five studies were classified as group A, indicating they had an aim to study sex differences in MT utilisation. One study was classified as group B, 6 as group C and 4 as group D. The full results of quality scoring and the classification systems can be found in supplementary material.

### Primary Outcome

One study^30^ reported the unadjusted OR for sex difference of thrombectomy use. Thus, we calculated the unadjusted OR for the remaining studies from the raw data provided. There was statistically significant heterogeneity between studies (Q=1043.13 p<0.0001, I^2^=98.56%), therefore a random effects model was used for the meta-analysis. The random effects summary unadjusted OR was 0.931 (95% CI 0.834-1.040, p=0.206), indicating women had lower odds of receiving MT but the difference was not statistically significant (Figure 2). A post-hoc decision was made to perform a sensitivity analysis; primary outcome was assessed using a fixed effect model: the summary unadjusted OR was 1.028 (95% CI 1.017-1.040, p<0.001), indicating that men are less likely to receive MT.

**Figure 2:**
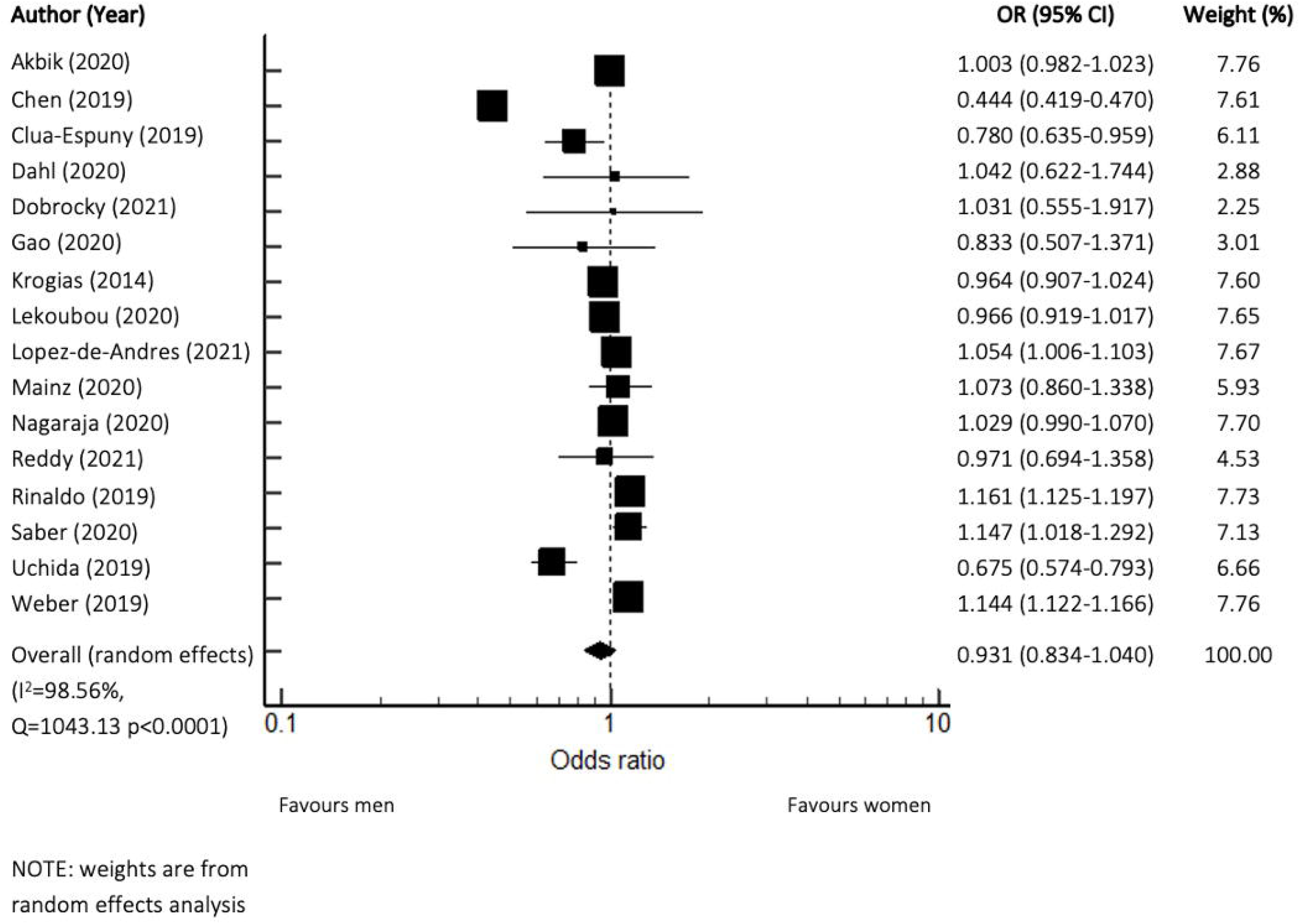
Forest Plot of unadjusted OR of MT use in women compared to men for all AIS admissions n=16 studies, CI = confidence interval

### Studies Providing Data on Adjusted OR

Only three studies^22,28,33^ (n=2464,979) provided data on adjusted OR for sex difference of thrombectomy use. The summary unadjusted OR was 0.66 (95% CI 0.37-1.18) with substantial heterogeneity (Q p<0.0001, I^2^=99.50%). The Individual factors that were adjusted for by the papers were as follows: one study did not report which factors they adjusted for^28^ and from the remaining two papers^22,33^, the only commonly adjusted for factor included age (table 2) A full list of adjustments can be found in table 2.

**Table 1:** Characteristics of the 16 included studies^16,17,21-34^

| Authors | Year of Publication | Country | Study Design | Time | Total number of AIS | Female MT treated cases, n | Female denominator, n | Female treatment rate, % | Male MT treated cases, n | Male denominator, n | Male treatment rate, % |
| --- | --- | --- | --- | --- | --- | --- | --- | --- | --- | --- | --- |
| Akbik et al. <sup>21</sup> | 2020 | United States | Registry-based | 2008-2018 | 267956 | 21651 | 134371 | 16.11 | 21479 | 133585 | 16.08 |
| Chen et al. <sup>22</sup> | 2019 | China | Administrative data | 2015 | 1443894 | 1512 | 583551 | 0.26 | 4966 | 853552 | 0.58 |
| Clua-Espuny et al. <sup>23</sup> | 2019 | Spain | Registry-based | 2011-2012 | 14368 | 165 | 7170 | 2.3 | 211 | 7198 | 2.93 |
| Dahl et al. <sup>24</sup> | 2020 | Sweden | Registry-based | 2014 | 1226 | 29 | 571 | 5.09 | 32 | 655 | 4.89 |
| Dobrocky et al. <sup>25</sup> | 2021 | Switzerland | Hospital-based | 2005-2020 | 169 | 33 | 65 | 50.77 | 52 | 104 | 50 |
| Gao et al. <sup>26</sup> | 2020 | Canada | Hospital-based | 2017-2018 | 256 | 51 | 130 | 39.2 | 55 | 126 | 43.7 |
| Krogias et al. <sup>27</sup> | 2014 | Germany | Administrative data | 2008 + 2012 | 458753 | 2101 | 233422 | 0.9 | 2103 | 225331 | 0.93 |
| Lekoubou et al. <sup>28</sup> | 2020 | United States | Administrative data | 2006-2014 | 1025477 | 3012 | 519310 | 0.58 | 3037 | 506167 | 0.6 |
| Lopez-de-Andres et al. <sup>29</sup> | 2021 | Spain | Administrative data | 2016-2018 | 172255 | 3602 | 79731 | 4.5 | 3976 | 92524 | 4.3 |
| Mainz et al. <sup>30</sup> | 2020 | Denmark | Registry-based | 2016-2017 | 5356 | 152 | 2308 | 6.59 | 188 | 3048 | 6.17 |
| Nagaraja et al. <sup>16</sup> | 2020 | United States | Administrative data | 2016 | 468630 | 5470 | 237510 | 2.3 | 5175 | 231120 | 2.2 |
| Reddy et al. <sup>31</sup> | 2021 | United States | Registry-based | 2015-2018 | 3873 | 71 | 1964 | 3.62 | 71 | 1909 | 3.72 |
| Rinaldo et al. <sup>32</sup> | 2019 | United States | Administrative data | 2016-2018 | 206853 | 8918 | 99258 | 8.98 | 8433 | 107595 | 7.84 |
| Saber et al. <sup>17</sup> | 2020 | United States | Administrative data | 2016-2017 | 103765 | 555 | 49081 | 1.1 | 540 | 54684 | 0.99 |
| Uchida et al. <sup>33</sup> | 2019 | Japan | Registry-based | 2014-2016 | 2399 | 521 | 1087 | 47.9 | 757 | 1312 | 57.7 |
| Weber et al. | 2019 | Germany | Administrative data | 2013-2017 | 1112570 | 21995 | 541169 | 4.06 | 20407 | 571401 | 3.57 |

**Table 2:** Variables adjusted for in individual studies included in the meta-analysis of adjusted Ors

| Author<br>(Year) | Unadjusted OR<br>(95% CI)<br>(Calculated) | Adjusted OR<br>(95% CI) | Variables adjusted for |
| --- | --- | --- | --- |
| Chen<br>(2019) | 0.44 (0.42-0.47) | 2 (1.89 to 2.13) | Patient characteristics: age, sex, medical insurance status, source of hospital admission, diagnostic correctness and CCI score. Hospital characteristics: hospital level |
| Lekoubou<br>(2020) | 0.97 (0.92-1.02) | 1.12 (1.05-1.18) | Not reported |
| Uchida<br>(2019) | 0.68 (0.57-0.79) | 0.71 (0.59-0.86) | Registration period, age, NIHSS score, Alberta stroke program early CT score, creatinine value, location of main occlusion, vessels involved in main occlusion, presence of multiple occlusions, cause of stroke, history of stroke, time since symptom onset to hospital, door use of IV thrombolysis |

### Subgroup Analysis

#### Geographic Region

There were two studies from Asia (n=1,439,502), seven from North America (n=2,076,810) and seven from Europe (n=1,764,697). The summary unadjusted OR for the Asian studies was 0.543 (95% CI 0.360-0.819, p=0.004) with significant heterogeneity (Q=22.91 p<0.0001, I^2^=95.63%) indicating that women were less likely to receive MT. The summary unadjusted OR for the North American studies was 1.045 (95% CI 0.975-1.121, p=0.211) with significant heterogeneity (Q=71.98 p<0.0001, I^2^=91.66%). The summary unadjusted OR for the European studies was 1.021 (95% CI 0.933-1.117, p=0.650) with significant heterogeneity (Q=46.15 p<0.0001, I^2^=87.00%).

#### Study Design

Studies were either hospital-based (n=425), registry-based (n=295,178) or administrative data (n=4,985,406). The summary unadjusted OR for the hospital-based was 0.906 (95% CI 0.615-1.336, p=0.618) with no significant heterogeneity (Q=0.28 p=0.600, I^2^=0.00%).The summary unadjusted OR for the registry based studies was 0.895 (95% CI 0.751-1.066, p=0.213) with significant heterogeneity (Q=28.49 p=<0.0001, I^2^=82.45%). The summary unadjusted OR for the administrative data studies was 0.953 (95% CI 0.804-1.130, p=0.581) with significant heterogeneity (Q=993.52 p<0.0001, I^2^=99.30%).

#### Quality Score

Studies were grouped based on quality score: score of five or less (n=684,272) and six or more (n=4,596,737). The summary unadjusted OR for the studies with quality score 5 or lower was 0.972 (95% CI 0.838-1.128, p=0.711) with significant heterogeneity (Q=41.77 p<0.0001, I^2^=88.03%). The summary unadjusted OR for the studies with quality score 6 or higher was 0.919 (95% CI 0.794-1.063, p=0.255) with significant heterogeneity (Q=972.75 p<0.0001, I^2^=99.07%).

#### Specific focus of individual studies

The summary unadjusted OR for the 5 Group A studies (n=2,614,432) was 0.978 (95% CI 0.878-1.090, p=0.687) with significant heterogeneity (Q=87.22 p<0.0001, I^2^=95.41%). The unadjusted OR for the 1 Group B study (n=14,368) was 0.780 (95% CI 0.635-0.959). The summary unadjusted OR for the 6 Group C studies (n=2,016,102) was 0.886 (95% CI 0.663-1.182, p=0.410) with significant heterogeneity (Q=852.11 p<0.0001, I^2^=99.41%). The summary unadjusted OR for the 4 Group D studies (n=636,107) was 1.011 (95% CI 0.946-1.080, p=0.753) with no significant heterogeneity (Q=5.31 p=0.151, I^2^=43.49%).

## Discussion

This meta-analysis found 16 studies, reported on over 5 million patients, from around the world reporting sex-specific data on MT utilisation. There was substantial variation in methodology between studies; including the study design (hospital-based, registry-based or administrative), length of study period (range from 12 months to 15 years) and the inclusion of out-of-hospital strokes. Our meta-analysis showed no difference in usage between me and women overall, though in our pre-specified subgroup analysis, thrombectomy use was less common in women in Asia.

We had wanted to perform separate analyses after adjusting for factors other than sex that might have influenced thrombectomy use but only three studies reported adjusted odds ratios. Data on time between symptom onset and presentation were not reported in the studies included in our meta-analysis, and so we were unable to adjust for this. Ideally future studies should provide both unadjusted odds ratios and also odds ratios corrected for the other key variables that might influence decisions about thrombectomy including age, comorbidity and time between stroke symptoms and presentation to hospital.

During analysis, a random effects model was used due to statistical heterogeneity between studies. Heterogeneity persisted throughout subgroup analysis and there were important differences in study methodology, including: type of strokes included, inclusion of both in-hospital and out-of-hospital strokes and data source. Although we had decided to include studies only in anterior circulation strokes, a small number of posterior circulation strokes may be included in analysis, as some studies used databases that did not clarify stroke subtype.

Our subgroup analysis found that women were less likely than men in the Asian studies to receive MT. This contrasts with results from the previous meta-analysis of sex differences in IVT treatment, which found no difference in IVT treatment between the sexes in Asian studies^5^.

Ideally, time of publication would have been carried out as subgroup analysis, separated by the publication of evidence for MT benefit. However, only one of the final studies was published before 2015. Although most included studies were published between 2019-2021, many used data collected before or during 2015. This meant we were unable to create subgroups of “Before 2015 publications” and “After 2015 publications”, and it remains unclear how the 2015 publications affected MT utilisation.

Sex and other protected characteristics (including, race and age) were reported poorly in the studies, with only three references discussing racial differences in stroke care^28,31,32^ (n= 1,236,203). Two of the included studies saw statistically significant differences in MT utilisation in Black and/or Hispanic patients when compared to White/non-Hispanic patients^28,31^. Another study included in our analysis contradicted these results and found no significant difference in MT utilisation based on race, however this study had a much smaller patient cohort^32^. The racial disparities may reflect complex factors, including geographical location and demographic differences^28^. Regardless, further studies and robust reporting are required to investigate potential differences.

A strength of this systematic review and meta-analysis was use of a published, peer-reviewed, methodology^5^. This provided a framework for analysis that allows results to be comparable to that of sex differences in IVT treatment. Additionally, two independent authors completed screening, data extraction and quality assessment, minimising selective outcome reporting^35^.

This study had several limitations. Firstly, due to limited resources, the bibliographies of the included studies were not systematically screened for further possible references. Additionally, MedCalc software^19^ had limited functions, preventing fuller analysis evaluating the effect that adjustment for confounding factors had on summary ORs. Multiple studies used national databases and International Classification of Disease (ICD) 10 codes, both of which have inherent issues: administrative data relies on human input with margin for error, and there is a possibility that data are not reported, including stroke location or subtype, time from onset, stroke outcome. Missing data will differ between sources but causes variability between analysed studies. In general, there was a lack of reporting of sex differences in MT utilisation, meaning we had to calculate unadjusted ORs to complete analysis.

Two included studies^16,17^ (n=572,395) used the same database with an overlap of three months, but a post-hoc decision was made to include both studies in the final analysis as the overlap was short compared to the overall length of both studies (12 months^16^ and 15 months^17^). This potentially introduces bias to our analysis, as one included study^17^ showed that women were statistically significant more likely to receive thrombectomy, though the other study^16^ found no significant difference in thrombectomy use between men and women.

Our searches were performed just over a year ago and we are aware of more recent publications. A retrospecitive observational study published in 2022 from Korea (n=322,796) found that female patients were statistically significantly less likely to receive thrombectomy^36^. Another retrospective observational study published in 2022 from America (n=302,965) also found that, after statistical adjustment, women were less likely to receive thrombectomy than men^37^. Contrary to these results, a study from Spain published in 2021 (n=172,255) found that thrombectomy was more likely to be used in women (reference).

Although our systematic review found no difference in provision of thrombectomy for men and women, it is nevertheless important that ongoing audits collect and report data on equity of access to thrombectomy, according to sex, and ideally other protected characteristics. We are aware that some ongoing audits do not routinely report equity of access, regarding protected characteristics. E.g. Scottish Stroke Audit 2021 report^38^. The 2020 report included total number of female and male stroke cases, but no sex-specific treatment details^39^. The Sentinel Stroke National Audit Programme (SSNAP) Eighth Annual Report also lacked information regarding protected characteristics^41^, as did the 2020 Irish National Audit of Stroke report^42^. To fully investigate equity of AIS treatments, registry studies and audits should automatically report patient protected characteristics and collect and report data on factors that might confound the relationship between sex and thrombectomy use, so that future systematic reviews can adjust for relevant factors, including age and time since symptom onset to presentation at medical services. Making this data transparent would facilitate systematic research, allowing robust research into equity of stroke care.

## Conclusion

This meta-analysis found no strong evidence that men or women are more likely to be given MT, but there was heterogeneity between studies that has affected this analysis. A statistically significant sex difference in MT use was seen in Asian studies, with women less likely to be treated. Further research is required to assess if sex disparities in MT use exist, what factors may be affecting them, and to observe any differences over time. Comprehensive reporting of all protected characteristics in stroke studies and audit reports will aid the investigation and resolution of any inequities in stroke care.

## Data Availability

All data produced in the present study are available upon reasonable request to the authors.

