## Supplementary materials for "Are There Sex Differences in Thrombectomy Utilisation in Treatment of Acute Ischaemic Stroke? A Systematic Review and meta-analysis"

### Supplementary Material

### Contents

##### Figures


1. Figure e-1: page 21
2. Figure e-2: page 22
3. Figure e-3: page 23
4. Figure e-4: page 24


##### Tables


1. Table e-1: page 25
2. Table e-2: page 26

#### Supplemental Figures

Figure e-1:

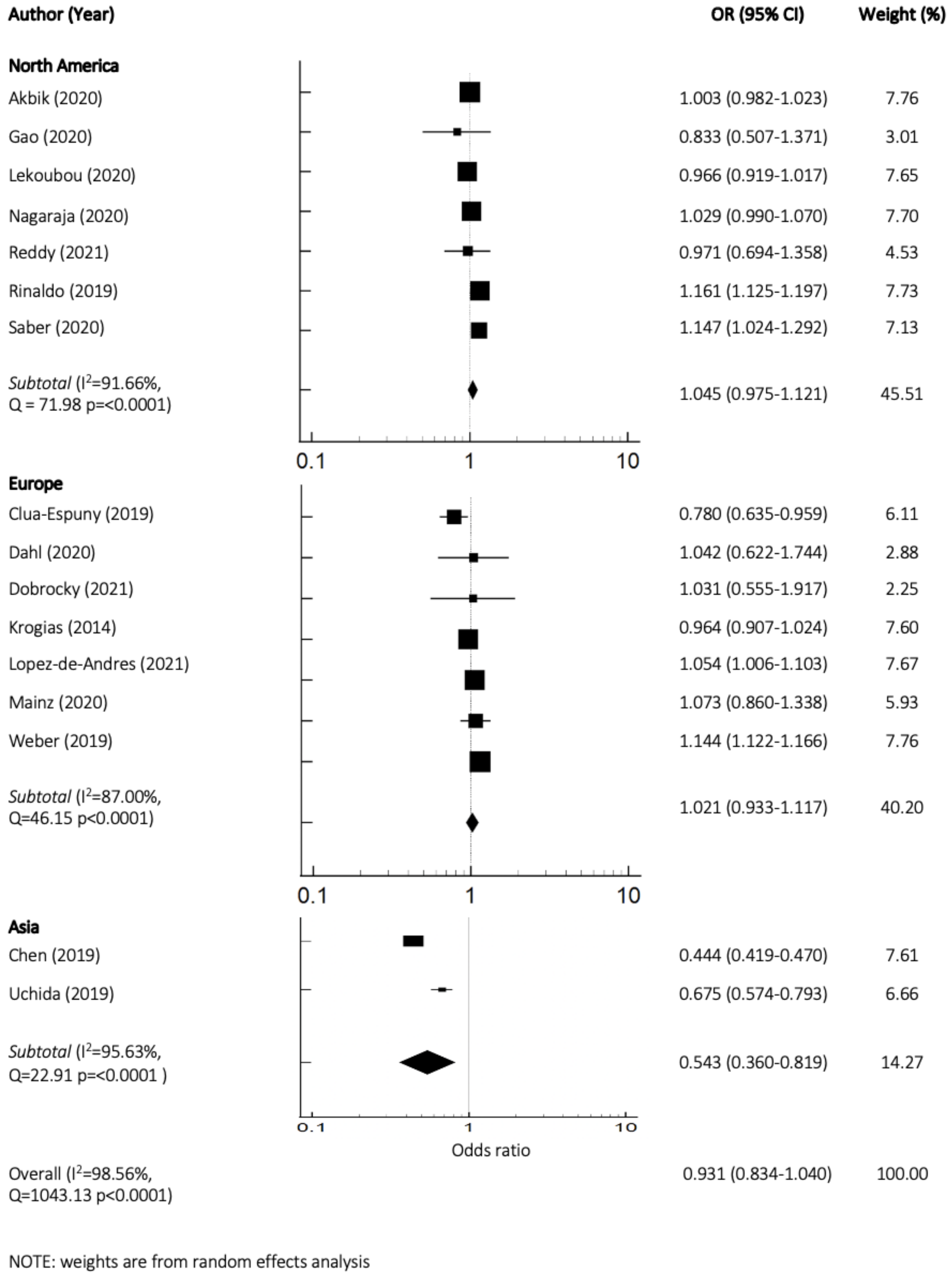


Sub-group analysis by geographic region (Asia, Europe, North America). Forest plots of the unadjusted OR of MT use in women compared to men in all acute ischemic stroke patients. Random effects model. (n= 16 studies)

Figure e-2:

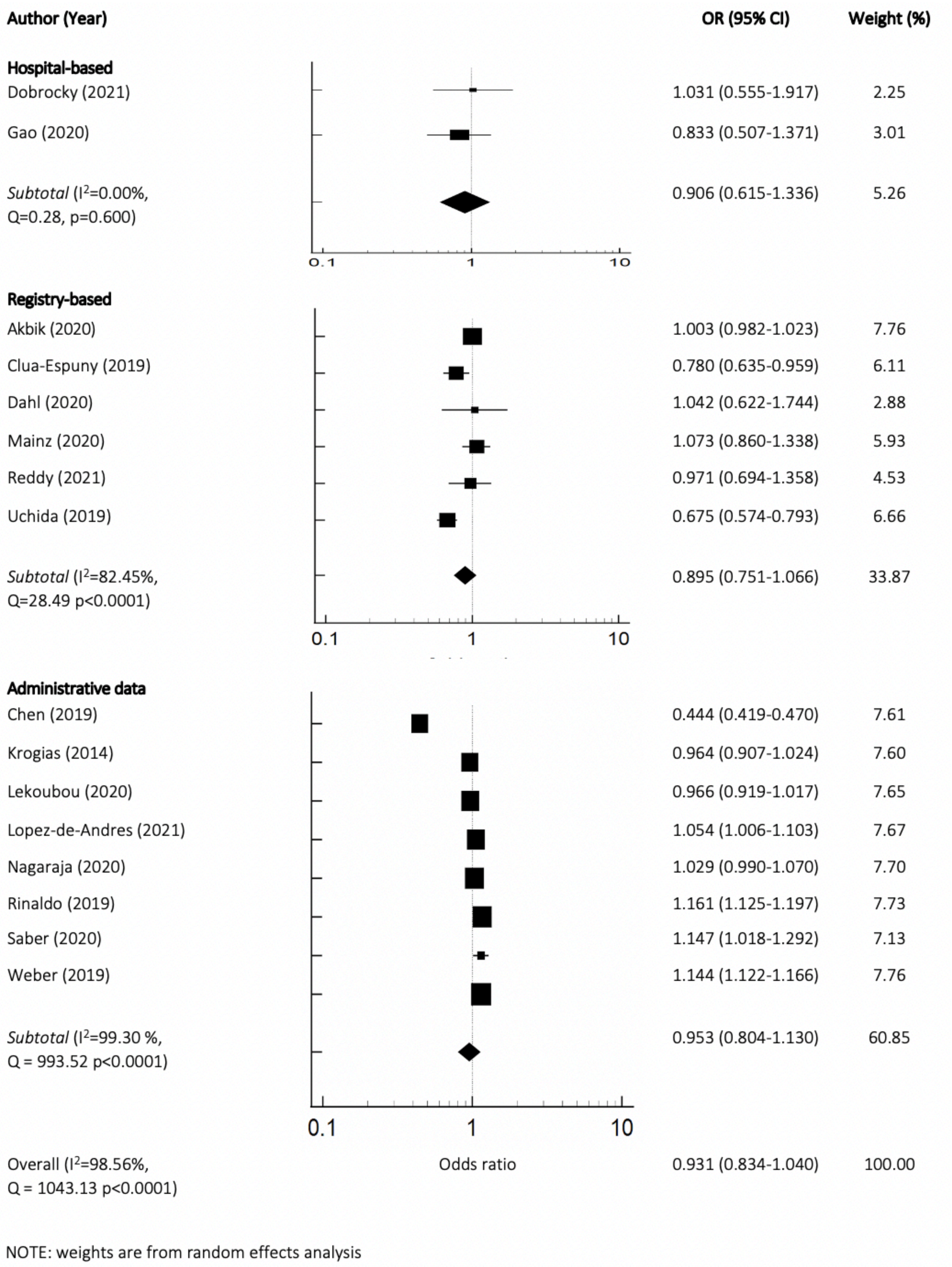


Sub-group analysis by study design (hospital based, administrative, registry). Forest plots of the unadjusted OR of MT use in women compared to men in all acute ischemic stroke patients. Random effects model. (n= 16 studies)

Figure e-3:

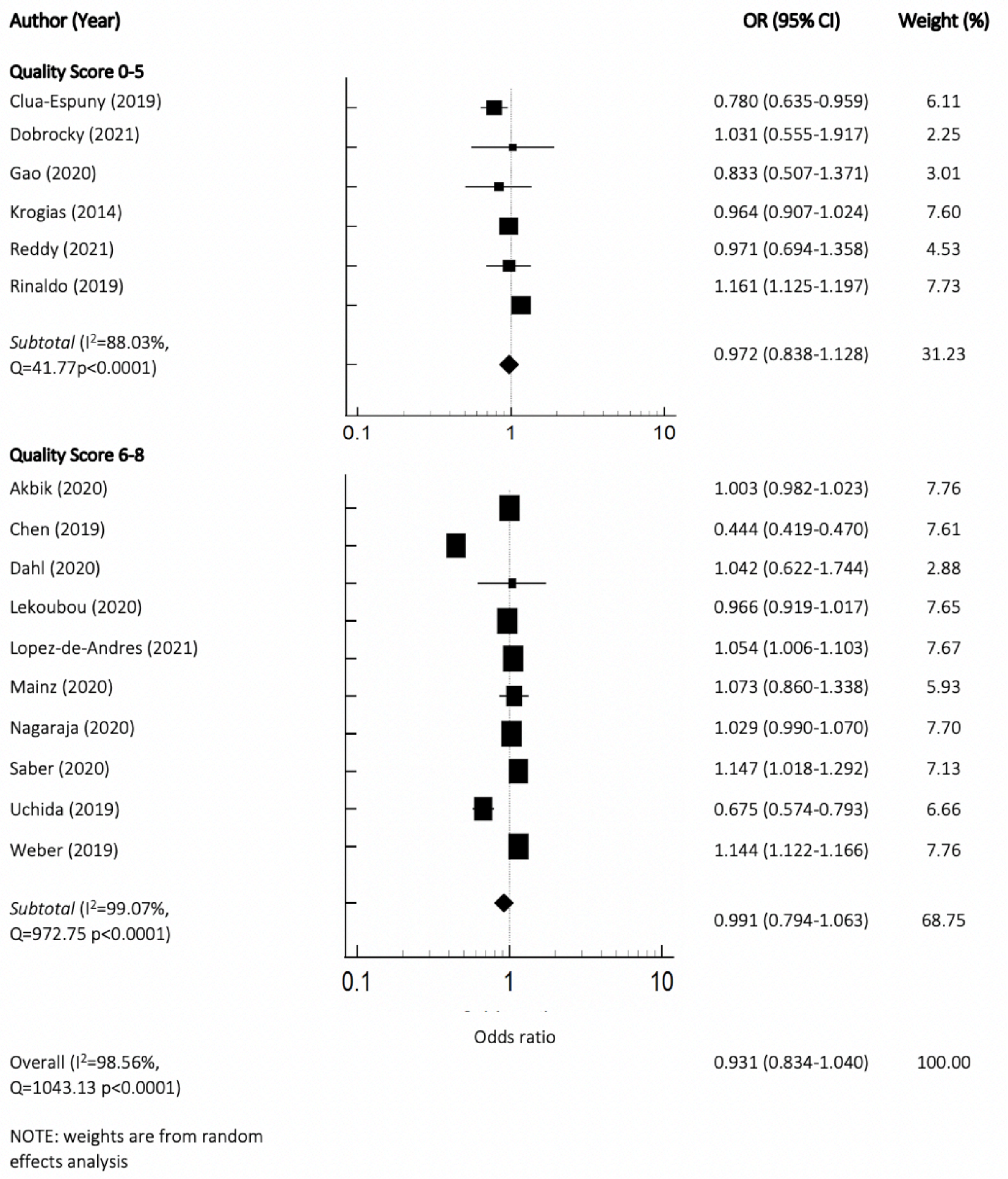


Sub-group analysis by quality score (0-8). Forest plots of the unadjusted OR of MT use in women compared to men in all acute ischemic stroke patients. Random effects model. (n= 16 studies)

Figure e-4:

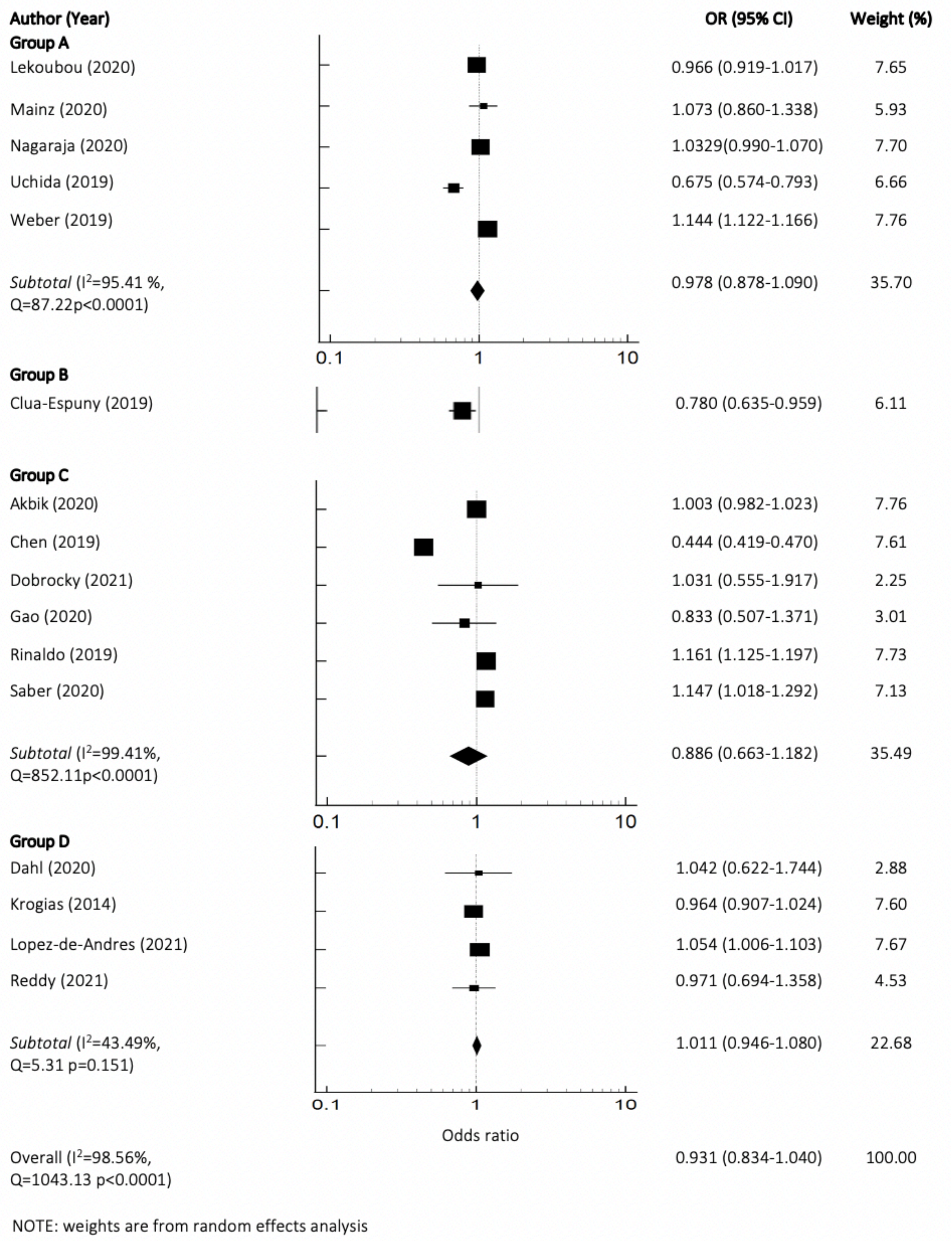


Sub-group analysis by primary study focus (A-D). Forest plots of the unadjusted OR of MT use in women compared to men in all acute ischemic stroke patients. Random effects model. (n= 16 studies)

### Supplemental Tables

Table e-1: Quality assessment scores and classifications for the 16 included studies

| Author (Year) | Representativeness ° | Exclusions † | Adjustment ‡ | Outcome § | Total | Primary focus |
|---|---|---|---|---|---|---|
| Akbik (2020) | 2 | 2 | 2 | 0 | 6 | C |
| Chen (2019) | 1 | 2 | 2 | 1 | 6 | C |
| Clua-Espuny (2019) | 1 | 2 | 1 | 1 | 5 | B |
| Dahl (2020) | 0 | 2 | 2 | 2 | 6 | D |
| Dobrocky (2021) | 0 | 2 | 2 | 0 | 4 | C |
| Gao (2020) | 1 | 0 | 2 | 2 | 5 | C |
| Krogias (2014) | 2 | 2 | 0 | 1 | 5 | D |
| Lekoubou (2020) | 2 | 2 | 1 | 1 | 6 | A |
| Lopez-de-Andres (2021) | 2 | 2 | 1 | 1 | 6 | D |
| Mainz (2020) | 2 | 2 | 2 | 1 | 6 | A |
| Nagaraja (2020) | 2 | 2 | 2 | 1 | 6 | A |
| Reddy (2021) | 1 | 2 | 0 | 2 | 5 | D |
| Rinaldo (2019) | 2 | 2 | 0 | 1 | 5 | C |
| Saber (2020) | 2 | 2 | 2 | 1 | 7 | C |
| Uchida (2019) | 2 | 2 | 2 | 0 | 6 | A |
| Weber (2019) | 2 | 2 | 0 | 1 | 6 | A |

For quality assessment, studies were rated in categories from 0-2 with 2 being the best and 0 the worst. Specific criteria for each score are detailed in the caption for table e-1. In terms of primary focus classification, the studies listed as "A" had a clear objective to study sex differences in thrombectomy use. Studies scored "B" examined sex differences in thrombectomy effectiveness but have no clear objective to study differences in use. Studies scored "C" focus on thrombectomy treatment more broadly with no objective to look at sex difference. Studies scored "D" focused on sex differences in acute stroke care with no objective to study thrombectomy use.

° The representativeness of the overall study population rated on a 0 to 2 point scale.**†** The number and impact of exclusions applied to initial patient cohort rated on a 0 to 2 point scale. **‡** The degree of adjustment for potential confounding variables that affect thrombectomy utilization rated on a 0 to 2 point scale. Studies that provided data on eligible treatment subgroups were not scored because statistical adjustment among patients already eligible for treatment is unnecessary. **§** The method by which outcome (MT treatment or not) was ascertained rated on a 0 to 2 point scale.

Table e-2: Example of quality assessments

| Author (Year) | Representativeness | Exclusions | Adjustment | Outcome | Total Score (out of 8) |
|---|---|---|---|---|---|
| Chen (2019) | 1 - This study used a subset of hospital type and assessed tertiary hospitals only. | 2 - The study population represented a consecutive cohort with exclusions that were less than 15%. | 2 - Adjusted for many patient characteristics (including age and sex) and hospital level. | 1 - The outcome was obtained through ICD-10 codes. | 6 |
| Lekoubou (2020) | 2 - This study used data from the National Inpatient Sample (NIS), which is a nationwide database in the USA. | 2 - The study population represented a consecutive cohort with exclusions that were less than 15%. | 1 - Adjustment reported but factors adjusted for are not reported. | 1 - The outcome was obtained through ICD-9 codes. | 6 |
| Weber (2019) | 2 - This study used nationwide data in Germany provided by the German Federal Statistic Office. | 2 - The study population represented a consecutive cohort with exclusions that were less than 15%. | 0 - Unclear if adjustment was carried out or not. ORs reported are not specified as unadjusted or adjusted. | 1 - The outcome was obtained through ICD-10 codes. | 6 |

Studies were assessed based on the representativeness of the overall study population, the number and impact of exclusions applied to the initial patient cohort, adjustment for potential confounding variables that affect MT utilization, and the method by which the outcome (MT treatment or not) was ascertained. With regard to representativeness, a score of *0* was given if the study included data from only one or two hospitals, *1* if the study was regional in scope and included multiple hospitals or studied a specific subset of hospitals (i.e., tertiary hospitals), and *2* if it was national in scope. For the number and impact of exclusions, a score of *0* was given if there were significant exclusions (>15%) from the starting cohort; *1* if the subjects under study comprised a random sample of a consecutive cohort, were taken only from stroke units, or represented cases selected as part of a randomized clinical trial; and *2* if it was a "gold standard" study (i.e., consecutive cohort with <15% exclusions). Regarding adjustment for confounding variables, a score of *0* was awarded for no adjustment, *1* for any adjustment at all, and *2* for full adjustment (e.g., age, sex, NIHSS score, etc.). Finally, for the method by which the outcome was ascertained, a score of *0* was given if there was no documentation of how the outcome was ascertained, *1* if it was done through billing codes, and *2* if it was done through the medical record.
